# Impact of environmental factors in predicting daily severity scores of atopic dermatitis

**DOI:** 10.1101/2020.10.27.20220947

**Authors:** Guillem Hurault, Valentin Delorieux, Young-Min Kim, Kangmo Ahn, Hywel C. Williams, Reiko J. Tanaka

**Affiliations:** Department of Bioengineering, Imperial College London, UK; Department of Pediatrics, Samsung Medical Center, Sungkyunkwan University School of Medicine; Environmental Health Centre for Atopic diseases, Samsung Medical Center, Korea; Centre of Evidence-Based Dermatology, University of Nottingham, UK

## Abstract

**Background:** Atopic dermatitis (AD) is a chronic inflammatory skin disease that affects 20% of children worldwide. Although environmental factors including weather and air pollutants have been shown to be associated with AD symptoms, the time-dependent nature of such a relationship has not been adequately investigated.

**Objective:** This paper aims to assess the short-term impact of weather and air pollutants on AD severity scores.

**Methods:** Using longitudinal data from a published panel study of 177 paediatric patients followed up for 17 months, we developed statistical machine learning models to predict daily AD severity scores for individual study participants. Exposures consisted of daily meteorological variables and concentrations of air pollutants and outcomes were daily recordings of scores for six AD signs. We developed a mixed effect autoregressive ordinal logistic regression model, validated it in a forward-chaining setting, and evaluated the effects of the environmental factors on the predictive performance.

**Results:** Our model outperformed benchmark models for daily prediction of the AD severity scores. The predictive performance of AD severity scores was not improved by the addition of measured environmental factors. Any potential short-term influence of environmental exposures on AD severity scores was outweighed by the underlying persistence of preceding scores.

**Conclusions:** Our data does not offer enough evidence to support a claim that AD symptoms are associated with weather or air pollutants on a short-term basis. Inferences about the magnitude of the effect of environmental factors require consideration of the time-dependence of the AD severity scores.

## INTRODUCTION

Atopic dermatitis (AD, also called eczema) is a chronic inflammatory skin disease characterised by inflammatory flares as well as dry and itchy skin [1]. AD patients often suffer from symptoms that fluctuate every day, resulting in a decreased quality of life due to the unforeseeable nature of the symptoms. AD affects almost 20% of the paediatric population worldwide and the prevalence of AD in children is still increasing globally [2]. The rising prevalence of AD coincides with increased urbanization and industrialization worldwide [3], and the assessment of the effects of environmental factors on AD has gained a growing importance.

AD pathophysiology is considered to be affected by external environmental factors, such as air pollution from particulates, ultraviolet radiation, temperature and humidity - collectively known as the skin exposome [4] [5]. Environmental factors have been shown to be associated with AD development and aggravation [6] [7], as well as other aspects of AD including barrier dysfunction [8] or care visits [9]. Prior studies investigated whether environmental factors were associated with the current AD severity [10] [11] [12] [13], but none have considered the dynamic nature of the severity nor have they investigated whether the future AD severity can be predicted by environmental factors. Despite this evidence gap, a profusion of smartphone eczema apps have emerged offering to track disease severity and environmental factors with bold claims of being able to predict AD flares [14].

We have recently developed statistical machine learning models to predict AD severity scores on a daily basis at an individual level [15]. The models demonstrated that it was possible to decipher much of the apparent unpredictable dynamics of AD severity scores from each patient’s longitudinal data. The models investigated the effects of age, filaggrin mutations and the treatments used, such as calcineurin inhibitors and corticosteroids, on daily change of AD severity scores. However, environmental stressors were only modelled as latent variables due to the lack of availability of such data in the training datasets.

In this paper, we aim to assess the impact of environmental factors in *predicting future AD symptoms*. We developed a statistical machine learning model to predict daily AD severity scores for individual patients using a longitudinal dataset with high quality environmental and AD symptom data. We used that model to evaluate whether environmental factors including weather and air pollutants are important determinants in predicting the next day’s AD scores from today’s scores.

## METHODS

### Data

We used the longitudinal data from a published panel study [10] that investigated the short-term impact of environmental factors on AD symptoms in Seoul, South Korea. The cohort included 177 Korean paediatric patients (67 girls and 110 boys) aged five or younger (average age of 2.0 years old, SD = 1.6) with mild to severe AD (mean SCORAD at enrolment of 31.1, SD = 12.8). The data contained the daily recording of the atopic dermatitis symptom score (ADSS) [16] over 17 months (Figs 1 and S1). ADSS is a sum of scores for six AD signs (dryness, edema, itching, oozing, redness and sleep disturbance), each on a discrete scale from 0 (none) to 4 (severe). In this study, we used the six AD sign scores, rather than their sum (ADSS), to extract more information from the data. 18.9% of the daily AD sign scores were missing. We removed five patients with less than ten daily observations, resulting in a total of 34921 patient-day observations.

**Figure 1:**
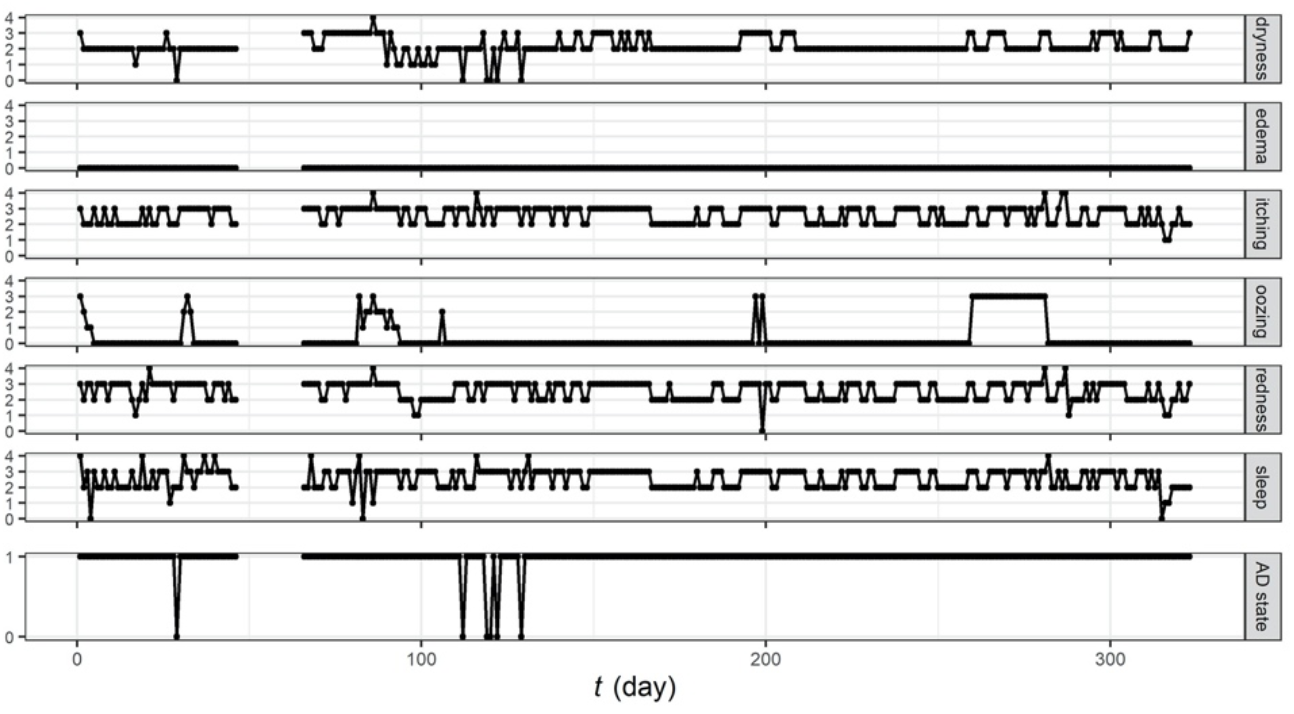
Example trajectories of the six AD sign scores and the derived AD symptom state for a representative patient.

The use of topical corticosteroids (binary) was recorded daily. Weather variables (mean temperature, relative humidity, total rainfall, diurnal temperature range) and the concentration of air pollutants (PM10, NO2, O3) were collected daily for each patient. A binary AD symptom state was derived in [10] from the sign scores: the state was 1 when the sum of itching and sleep disturbance scores was greater than or equal to 2 and the scores of at least two of redness, dryness, edema, or oozing were non-zero, and the state was 0 otherwise (Fig 1).

### Mixed effect autoregressive ordinal logistic regression model

We developed a mixed effect autoregressive ordinal logistic regression model to predict the patient-dependent dynamics for each of the six AD sign scores. The model is described by

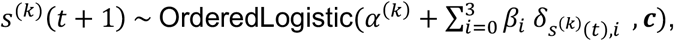

where *s*^(*k*)^(*t*) is a sign score for the *k*-th patient at day *t, α*^(*k*)^ is the patient-dependent intercept (the random effect), *β*_*i*_’s are the regression coefficients, *δ*_*x,y*_ is the Kronecker delta, and ***c*** is the vector of cut-off values of the ordered logistic distribution (details in the SI Text). The linear predictor, 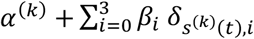, corresponds to the location parameter of the ordered logistic distribution. We also considered a model with all covariates of interest (the environmental variables and TCS usage at *t* for evaluation of the impact of environmental factors) included in the linear predictor and models with one covariate each. Cross-correlation analysis did not support the inclusion of higher order time lags for sign scores or covariates in the model. The models were fitted, using the “lme4” package in R, to pairs of successive scores (*s*^(*k*)^(*t* + 1), *s*^(*k*)^(*t*)). Pairs with at least one missing value were removed from the training set.

### Model validation

We evaluated the predictive performance of our models in a forward-chaining setting where the models were trained with the first day’s data and tested on the second day’s data, then re-trained on the first two days’ data and tested on the third day’s data, and so on. The performance of predicting AD sign scores was quantified by the ranked probability score (RPS), a proper scoring rule for ordinal probabilistic forecasts. The performance of predicting binary AD symptom states was evaluated with the Brier score.

We compared the performance of our models to that of two benchmark models: the uniform forecast model that predicts each of the five possible outcomes of a sign score with the probability of 1/5, and the historical forecast model where the probability of each possible outcome is equal to its occurrence in the patient’s training data. We also compared our models to the logistic regression model proposed in [10] for the prediction of AD symptom states.

## RESULTS

### Model validation

We trained each of the six mixed effect autoregressive logistic regression models without covariates, where each model was developed for prediction of one of the AD sign scores. The models learnt the patient-dependent dynamics of the sign scores as more data came in and outperformed the benchmark models in predicting the next day’s score for all AD signs (Fig 2). The performance of the benchmark models varied across signs, confirming that the scores of some signs are more imbalanced than others (Fig S1) and easier to predict. For instance, the historical forecast model (and our model) achieved an almost perfect prediction for edema, for which the outcome is 0 for nearly 90% of the time. For other signs, such as dryness, the RPS of our model was about 60% lower (i.e. achieved a better performance) than that of the historical forecast model after 200 days of training.

**Figure 2:**
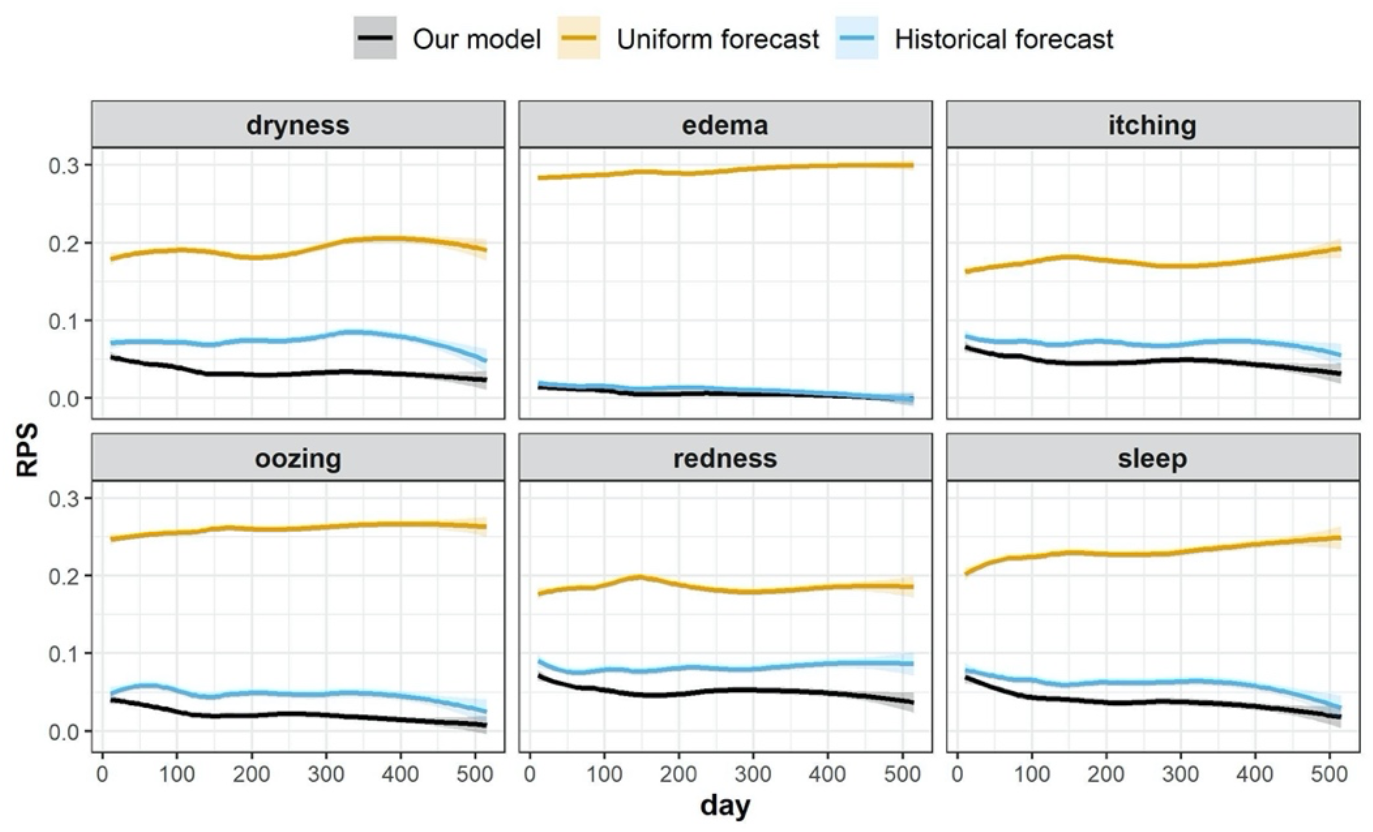
Comparison of the predictive performance of our model (the mixed effect autoregressive ordinal logistic regression without covariates) to that of the uniform forecast and the historical forecast models, for prediction of each of the six AD signs. The performance of predicting AD sign scores is measured by the RPS (the lower RPS indicates the better predictive performance). Learning curves were obtained using locally weighted scatterplot smoothing (LOWESS). Shaded areas correspond to ± 1.96 standard error.

We derived a prediction for the binary AD symptom state from the six mixed effect autoregressive logistic regression models for AD sign scores (Fig S2), assuming their predictions are independent random variables. Our model outperformed the two benchmark models and the logistic regression model proposed in [10]. The Brier score of our model was about 40% lower (i.e. achieved a better performance) than the logistic regression model, which performed similarly to the historical forecast.

### Effect of environmental factors on the model’s predictions

To assess the effects of exogenous factors (weather, air pollution, TCS usage) on the prediction of AD sign scores, we computed the pairwise difference in the RPS between the model without covariates, the models with a single covariate, and the model with all covariates (Fig 3).

**Figure 3:**
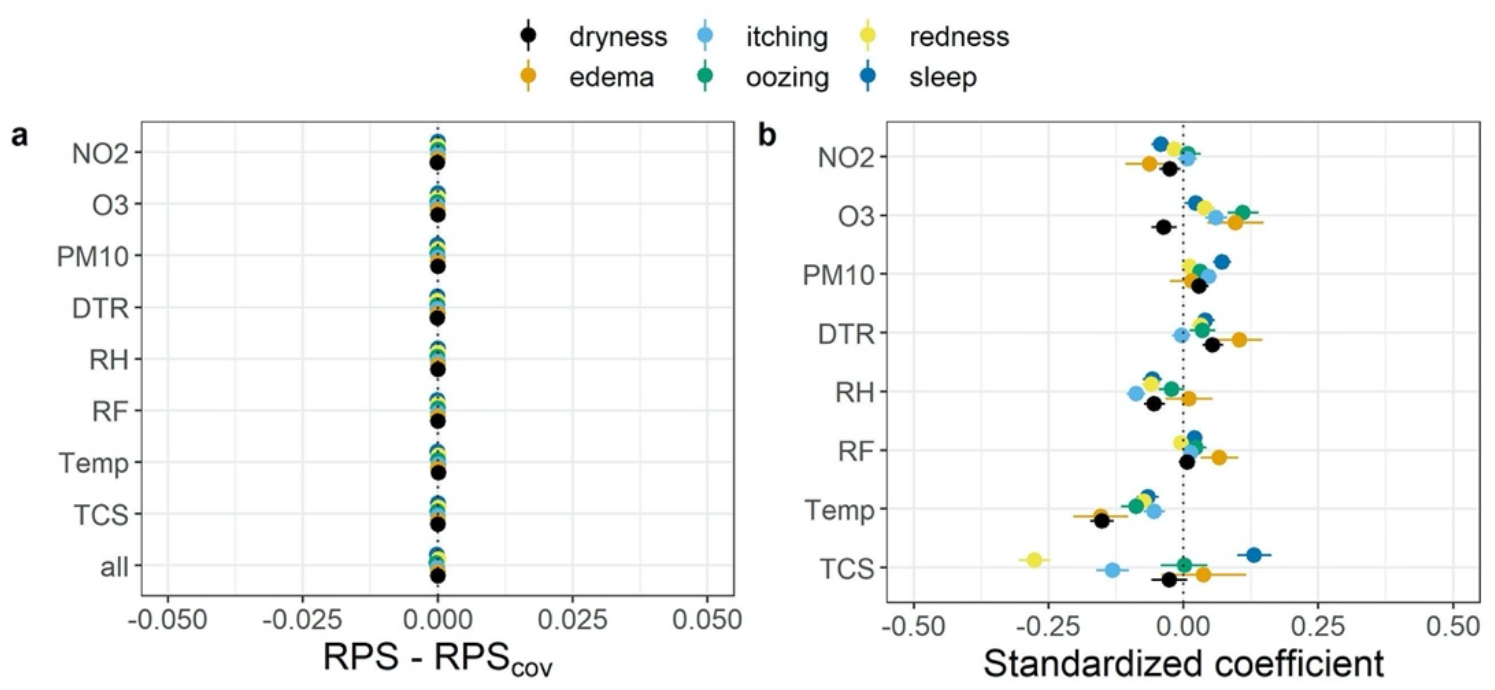
Effects of environmental factors (mean temperature (Temp), relative humidity (RH), total rainfall (RF), diurnal temperature range (DTR), and the concentration of air pollutants (PM10, NO2, O3)) and treatment usage (topical corticosteroids (TCS)) on AD sign score prediction. (a) The pairwise difference in predictive performance between the model without covariate (RPS) and the model with covariates (single or all, *RPS*_*cov*_). *RPS*−*RPS*_*cov*_> 0 indicates that the model with covariates has a higher predictive performance. (b) The coefficients for the covariates in the single-covariate models (± *SE*). A positive coefficient means that an increase in the covariate is associated with a higher probability for more severe outcomes.

No evidence was found to support that the inclusion of exogenous factors improved the predictive performance of the model for all signs (Fig 3a). Even though some of the coefficients associated with the covariates have confidence bounds that do not cross 0, all of them were small in magnitude accounting for approximately 1% of the linear predictor (Fig 3b). These small coefficients result in the lack of a noticeable improvement in the predictive performance of the model by addition of the covariates.

## DISCUSSION

We developed a mixed effect autoregressive ordinal logistic regression model that can predict the next day’s AD severity scores, using the longitudinal data from a published panel study [10]. Our model outperformed two benchmark models for the prediction of AD sign scores (Fig 2), and outperformed the benchmark models and the logistic regression model for the prediction of an AD symptom state proposed in [10] (Fig S2). Despite development of such a model, we showed that inclusion of environmental factors did not improve the predictive performance of the model (Fig 3).

Our results from a comprehensive dataset of South Korean children does not present any convincing evidence to support a claim that AD symptoms were associated with weather or air pollutants on a short-term basis. The short-term influence of the environmental factors on AD sign scores was outweighed by the previous scores’ persistence, and the next day’s score for a patient is more accurately predicted using the patient’s today’s score than using environmental data. Neglecting the time-dependence of the AD symptoms severity scores as in previous studies [9] [11] [12] [13] may misguide inferences about the effect size of environmental associations. The extent to which AD severity can be predicted from the measurement of environmental factors remains unclear. Our results throw serious doubts into the claim of many AD apps that purport to use real time environmental measures to inform AD users when their AD symptoms are likely to flare.

It is possible that other “internal” factors such as the development of skin autoimmunity may be more important than external factors in determining disease fluctuations over time [17]. It is also important to state that factors that determine disease incidence may be different from those that determine disease chronicity, so it is still possible that environmental factors may be more predictive of the AD onset and long-term disease trajectories rather than short-term symptom fluctuations.

This study used the high-quality dataset on South Korean children with high rates of data completion. Modelling each of the six AD signs enabled to extract more information from the data and to generate predictions for any quantity of interest to the practitioner, be it ADSS or any combination of the sign scores. In terms of study limitations, the AD sign scores used in this study were obtained by subjective assessment by the patients (or their carers) on a discrete scale. Further investigation of the seemingly small effects of environmental factors on AD severity scores may benefit from more data or better quality data, for example by recording time-series of SCORAD (severity scoring of atopic dermatitis) or EASI (eczema area and severity index), or their self-assessed version, as they are more objective and may offer better responsiveness to environmental changes. However, dichotomisation of AD sign scores into a binary AD symptom state as proposed in [10] reduces the power of the analysis [18] and is not recommended. Our model might be improved by taking measurement errors into account using hidden Markov models or by modelling the correlations between AD signs in a multi-outcome regression. However, we believe the additional complexity in the model would only result in marginal improvements in the already solid predictive performance.

Whilst this study focused on the association between environmental factors and future AD severity scores, whether environmental factors *cause* a change in AD scores is of more interest for the AD community. Estimating the causal effect is challenging, as most causal inference methods assume the absence of unobserved confounders [19], an assumption that is deemed unrealistic. For example, “staying indoors” was not recorded in the original study [10] but could lead to reverse causation if patients decided to stay indoors during a pollution peak. Estimation of non-linear interactions may also be required, if patients react differently to environmental triggers depending on their severity: mild patients could be less sensitive than severe patients who may be subject to a “ceiling effect”. Constructing causal diagrams using specialist background knowledge could be a promising approach.

The methods presented in this study could be applied to other diseases, such as asthma, for which associations between environmental factors and asthma exacerbations are of interest.

## Data Availability

All the codes are available at https://github.com/VDelorieux/AD-environmental_factors.

## DATA AVAILABILITY

All the codes are available at https://github.com/VDelorieux/AD-environmental_factors.

## ACKNOWLEDGEMENTS

This study was funded by British Skin Foundation (005/R/18) and by the Environmental Health Center Project of the Ministry of Environment, Republic of Korea.

## Supporting Information Text

The probability mass function of the ordered logistic distribution for an outcome *y* ∈ {0,1,2,3,4} is parametrised by a location *η* and a vector of cut-off value, ***c*** = (*c*_0_ *c*_1_ *c*_2_ *c*_3_), *c*_0_ < *c*_1_ < *c*_2_ < *c*_3_, as

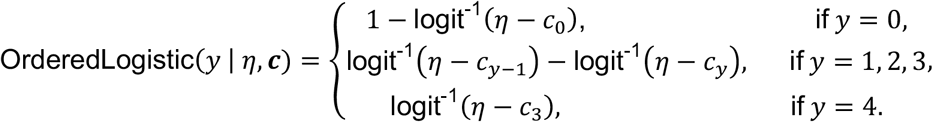

In practice, the logits can be obtained by jointly fitting the cumulative distribution, *P*(*y* ≤ 0), *P*(*y* ≤ 1), *P*(*y* ≤ 2) and *P*(*y* ≤ 3), with logistic regressions.

## Supporting Figures

**Figure S1:**
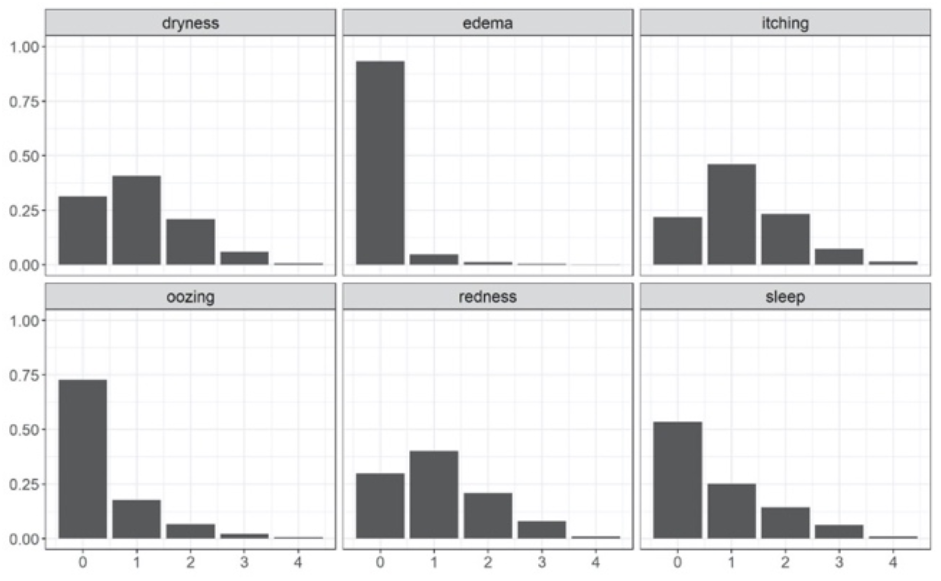
Distribution of the AD signs scores across time and patients.

**Figure S2:**
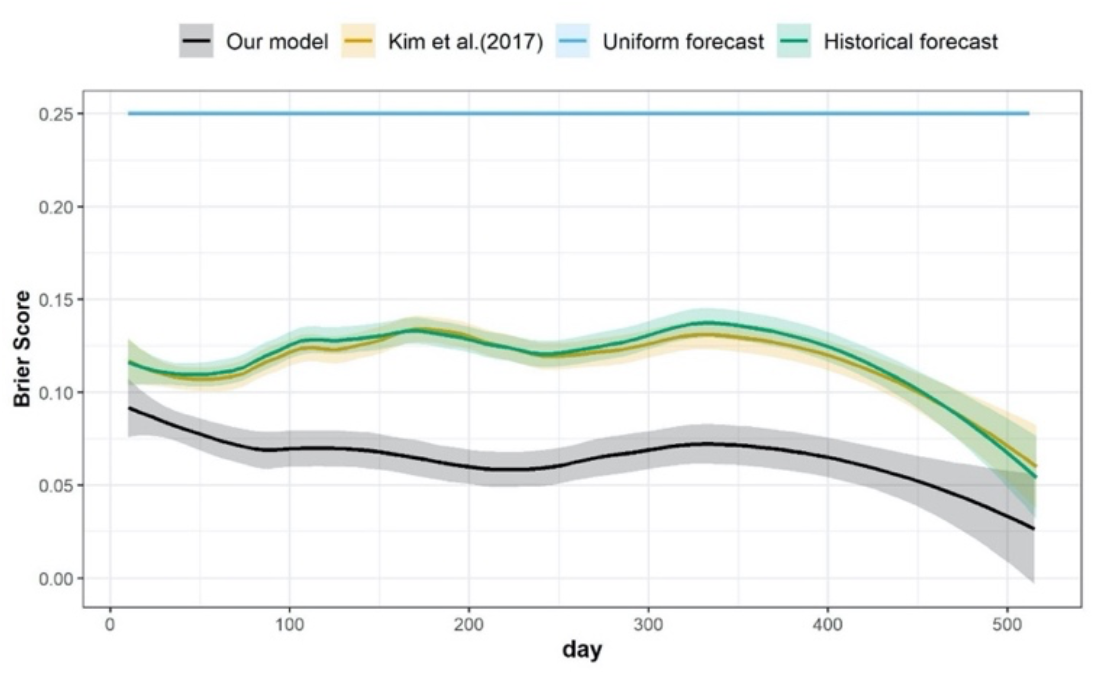
Comparison of the predictive performance for the models predicting the AD symptom state. Our model (without covariate and for which the prediction for the AD symptom state is derived from the predictions for each AD sign) is compared to the uniform and the historical forecast models, and the logistic regression model proposed in Kim et al. (2017)[10]. The performance is measured by the Brier score (the lower Brier score corresponds to the better predictive performance). Learning curves were obtained using LOWESS smoothing. Shaded areas correspond to ± 1.96 standard error.

## Notes

### Competing Interest Statement

The authors have declared no competing interest.

